# Affinity Proteomics for Saliva Biomarker Discovery Using High-Throughput Proximity Extension Assay

**DOI:** 10.64898/2026.03.13.26347316

**Authors:** Melissa M. Grant, Monique Stoffels, Matthias Born, Iain L. C. Chapple

**Author notes:** Correspondence: Melissa Grant, Periodontal Research Group, Dentistry, School of Health Sciences, College of Medicine and Health, University of Birmingham and Birmingham Dental Hospital (Birmingham Community Healthcare Trust), 5 Mill Pool Way, Edgbaston, Birmingham, B5 7EG.

## Abstract

Saliva offers a non⍰invasive, low⍰cost, and patient⍰friendly matrix for biomarker discovery. Affinity⍰based proteomic technologies such as the Proximity Extension Assay (PEA) are increasingly being adopted for large⍰scale biomarker studies, yet they remain underexplored in saliva. This study applied the Olink Explore High⍰Throughput (HT) PEA platform to profile approximately 5,400 proteins in saliva samples collected from donors representing periodontal health, gingivitis, advanced periodontitis (baseline and 3⍰months post⍰treatment), and edentulism.

Saliva from 68 donors was analysed, and all samples passed Olink’s quality⍰control procedures, with only 17 of 5,416 assays failing. Forty⍰one percent of proteins were detected above the limit of detection, demonstrating substantial assay sensitivity in this biofluid. Principal component analysis revealed clear compositional differences between clinical groups, with post⍰treatment periodontitis samples clustering more closely with health than baseline disease. Pairwise group comparisons identified hundreds of differentially abundant proteins, with consistently more proteins increased than decreased relative to health.

This study demonstrates, for the first time, that Olink HT can robustly measure thousands of proteins in saliva with high data quality and biologically meaningful discrimination between periodontal states. The platform’s minimal sample⍰volume requirements and scalability present strong potential for future saliva⍰based biomarker discovery and translational research.

## Introduction

Saliva is a biofluid rich in proteins and has emerged as an attractive biofluid for proteomic investigations into discovery and translational medical science due to its non-invasive collection, minimal risk, low cost, and high patient acceptability (1, 2). Unlike blood-based sampling, saliva collection does not require clinically trained personnel or venipuncture, thereby facilitating large-scale longitudinal studies, repeated sampling in vulnerable populations, and community-based screening. The salivary proteome reflects contributions from multiple sources, including major and minor salivary glands, gingival crevicular fluid, oral epithelial cells, and resident microbiota, creating a rich molecular landscape that can capture both local oral physiology (e.g., periodontal inflammation, caries risk, mucosal health) and systemic pathophysiology (e.g., metabolic, endocrine, immune dysregulation). In recent years, evidence has accumulated that salivary proteins and peptides can act as biomarkers of multiple biological processes. In 2022 we reported that saliva can be a source of biomarkers for periodontal diseases and that a small panel of biomarkers can distinguish between saliva donated by orally healthy individuals, and those with gingivitis or periodontitis (3).

Nevertheless, comprehensive profiling of the salivary proteome presents various analytical challenges (4). Protein concentrations in saliva are typically lower than in plasma, and the matrix contains abundant enzymes (including proteases), mucins, and variable levels of contaminants that can interfere with protein/peptide detection. The dynamic range of protein abundance in saliva is substantial (5), and many disease-relevant proteins are present at low concentrations, demanding highly sensitive assays with high specificity.

Discovery proteomic exploration of saliva has historically relied on mass spectrometry (MS), including data-dependent and data-independent acquisition strategies, often preceded by fractionation, depletion of abundant proteins, or enrichment of specific classes of molecules (6). MS-based methods offer unbiased and often untargeted discovery, the ability to identify novel peptides and post-translational modifications and continually improving depth of coverage. However, MS in saliva can encounter practical limitations: sample preparation can be complex; matrix components (e.g., mucins, proteolytic activity) can impair peptide detection; quantification at very low abundance is challenging; and throughput constraints may limit the number of samples and conditions profiled within realistic budgets and timelines.

Affinity-based proteomics offers a complementary route, prioritizing sensitivity and specificity through antibody- or binder-mediated recognition. Techniques such as multiplex immunoassays, aptamer-based assays, and proximity-based immunoassays have expanded the measurable protein space, particularly for low-abundance analytes that are difficult to quantify by MS in complex matrices (7). Crucially, these platforms can scale to thousands of targets in parallel and are amenable to the study of large cohorts, improving statistical power for biomarker discovery and evaluation. Yet, the application of truly large-scale affinity proteomics in saliva remains underexplored compared to plasma or serum.

Proximity Extension Assay (PEA) is an affinity-based proteomic technology that employs pairs of oligonucleotide-labelled antibodies recognizing distinct epitopes on the same target protein(8). Upon dual binding, the conjugated oligonucleotides are brought into close proximity, enabling a DNA polymerase–mediated extension or ligation event that forms a unique amplifiable sequence. Quantification is then achieved through highly sensitive nucleic acid amplification and detection, translating protein recognition into a digital nucleic acid signal. This dual-recognition architecture enhances specificity by requiring simultaneous binding of two independent probes, which reduces off-target signals. Signal amplification through DNA-based methods provides exceptional sensitivity, enabling detection of low-abundance proteins in challenging matrices such as saliva.

Here we apply PEA at large scale, profiling approximately 5,400 proteins to characterize the salivary proteome with breadth and depth. Such scale allows investigation of diverse biological processes, including innate and adaptive immunity, tissue remodelling, host-microbe interactions, metabolism, and intercellular communication. It also supports systems-level analyses, such as pathway enrichment, which can illuminate disease-relevant modules and candidate biomarker panels with potential translational utility.

## Materials and Methods

### Study population

Saliva samples were sourced from a previous collaboration between Philips Oral Healthcare and Newcastle University and The University of Birmingham. Participants were recruited to one of five groups, as defined in Table 1. All participants were recruited between 2009 and 2012. Population demographics can be found in Table 2.

**Table 1.** Definitions of participant groups.

| Group | Definition |
| --- | --- |
| Health | • No sites with interproximal attachment loss<br>• PD $\leq 3$ mm in all sites (but would allow up to four 4-mm pockets at distal of last standing molars)<br>• $\leq 10\%$ sites with mGI of $\geq 2.0$<br>• $< 10\%$ sites with BOP |
| Gingivitis | • No sites with interproximal attachment loss<br>• $> 30\%$ of sites with mGI $\geq 3.0$<br>• BOP scores $> 10\%$<br>• No sites with PD $> 4$ mm |
| Generalized Advanced periodontitis (Stage III/IV under 2018 classification) | • Interproximal PPD of $> 7$ mm (equating to approximately $\geq 5$ mm CAL) at $\geq 12$ teeth<br>• BOP scores of $> 30\%$ |
| Edentulous | • Completely edentulous for $> 1$ year with healthy oral tissues |

**Table 2.** Clinical data.

|  | **Health** | **Gingivitis** | **Advanced**<br>**periodontitis**<br>**(Stage III/IV)** | **Advanced**<br>**periodontitis**<br>**after treatment**<br>**(Stage III/IV)** | **Edentulous** |
| --- | --- | --- | --- | --- | --- |
| Number in group | 17 | 17 | 17 | 17 | 17 |
| Age (years)<br>mean (SD) | 35 (11.9) | 32.8 (9.7) | 50.8 (7.3) | 50.8 (7.3) | 68.2 (10.3) |
| Gender (%<br>female) | 55% | 35% | 53% | 53% | 53% |
| Probing<br>pocket depth<br>(mm) mean<br>(SD) | 1.4 (0.2) | 1.7 (0.2) | 3.7 (0.8) | 2.7 (0.7) | — |
| Clinical<br>attachment<br>level (mm)<br>mean (SD) | 0 | 0 | 4.9 (1.2) | 4.4 (1.4) | — |

### For the full cohort

140 medically healthy adult subjects (53% female) were recruited. The study was approved by County Durham & Tees Valley 1 NHS Research Ethics Committee, (ref. 09/H0905/49), and all volunteers provided written informed consent. 17 samples were selected per donor type.

Volunteers in all groups (defined in Table **1**) were examined and had biological samples collected and clinical indices examined to confirm their periodontal status. Clinical examination involved determination of clinical attachment level (CAL), probing pocket depths (PPD), bleeding on probing (BOP), modified gingival index GI (9) and plaque scores (10), using a UNC-PCP15 periodontal probe. Participants with gingivitis received standard oral hygiene instruction and were provided with a professional prophylaxis. Participants with periodontitis received non-surgical periodontal therapy by a dental hygienist until they achieved clinical outcomes that were consistent with successful periodontal therapy: endpoint of ≤4 sites with PD ≥5□mm, and ≤10% of sites with BOP (11). Periodontitis patients returned for two further oral hygiene instruction reinforcement visits at approximately monthly intervals, as part of their routine clinical care. Post-operative review and post-therapy biological sample collection were performed 3 months following treatment completion.

### Inclusion and exclusion criteria

Patients were excluded if they had less than 24 natural teeth (except edentulous patients), were current smokers, or had smoked up to within 5□years; wore removable dentures (partial or full) or bridges involving >4 teeth (except for the edentulous group); wore orthodontic appliances; were on long‐term antibiotic/anti‐inflammatory therapy or had taken antibiotic/anti‐inflammatory medication during the month prior to baseline assessment; were pregnant, breast feeding, or had medical/dental conditions incompatible with participation in study.

### Sample collection

Participants were asked to refrain from brushing, eating, or drinking (except water) for 2 h before sample collection. Samples were collected before oral examination and probing. Saliva production was stimulated using a sterilized marble.

### Olink analysis

Firalis Molecular Precision (France) provided a service to analyse the samples via the Olink Explore HT panels. Saliva (50ul per sample) was added to a 96 well plate and shipped on dry ice to Firalis, where they conducted the assays in accordance with Olink’s specifications and provided a full breakdown of all quality controls (QCs), and the resulting data, as normalised protein values (NPX), in an Excel spreadsheet. All samples were run undiluted (neat) in all blocks.

### Data analysis

R was used with the Olink Analyze package and further R functions in Bioconductor clusterProfiler.

## Results

Saliva was analysed from 68 donors with either health, gingivitis, advanced periodontitis or being edentulous. In addition, samples from the periodontitis donors were analysed 3-months following non-surgical periodontal treatment. All the saliva samples supplied for HT analysis passed the quality control (QC) standards. Of the 5416 assays (or proteins) only 17 did not pass the QC standards. As one plate was run the intra-assay coefficient of variance (%CV) was calculated: 10.6%. The CV was calculated for NPX of pre-selected assays with good performance in Olink Sample Controls, using the assumption of a log-normal distribution, in sample controls. Forty one percent of the assays had values above the limit of detection for NPX and 31% had values with >80% above the limit of detection (LOD). This is very similar to values calculated by Rooney et al for plasma proteins evaluated by Olink HT(12). On the subsequent analyses the evaluation is based on all the data including those below the LOD.

Principal component analysis (PCA) demonstrated a difference between the groups (figure 1A) with treated periodontitis samples being more similar compositionally to health or gingivitis samples than the baseline (untreated) periodontitis groups were. Using pairwise comparisons between all the groups and multiple comparison corrected nonparametric tests, it was possible to find multiple differences between the groups (Figure 1B-F and Table 3). When analysing contrasts between healthy donors and any other donor there were consistently more highly abundant than low abundancy proteins. These were plotted as a Venn diagram (Figure 1 G) and the more abundant proteins denoted as UP on the volcano plots were analyzed by gene ontology biological process for over enrichment (Figure 1 H).

**Figure 1.**
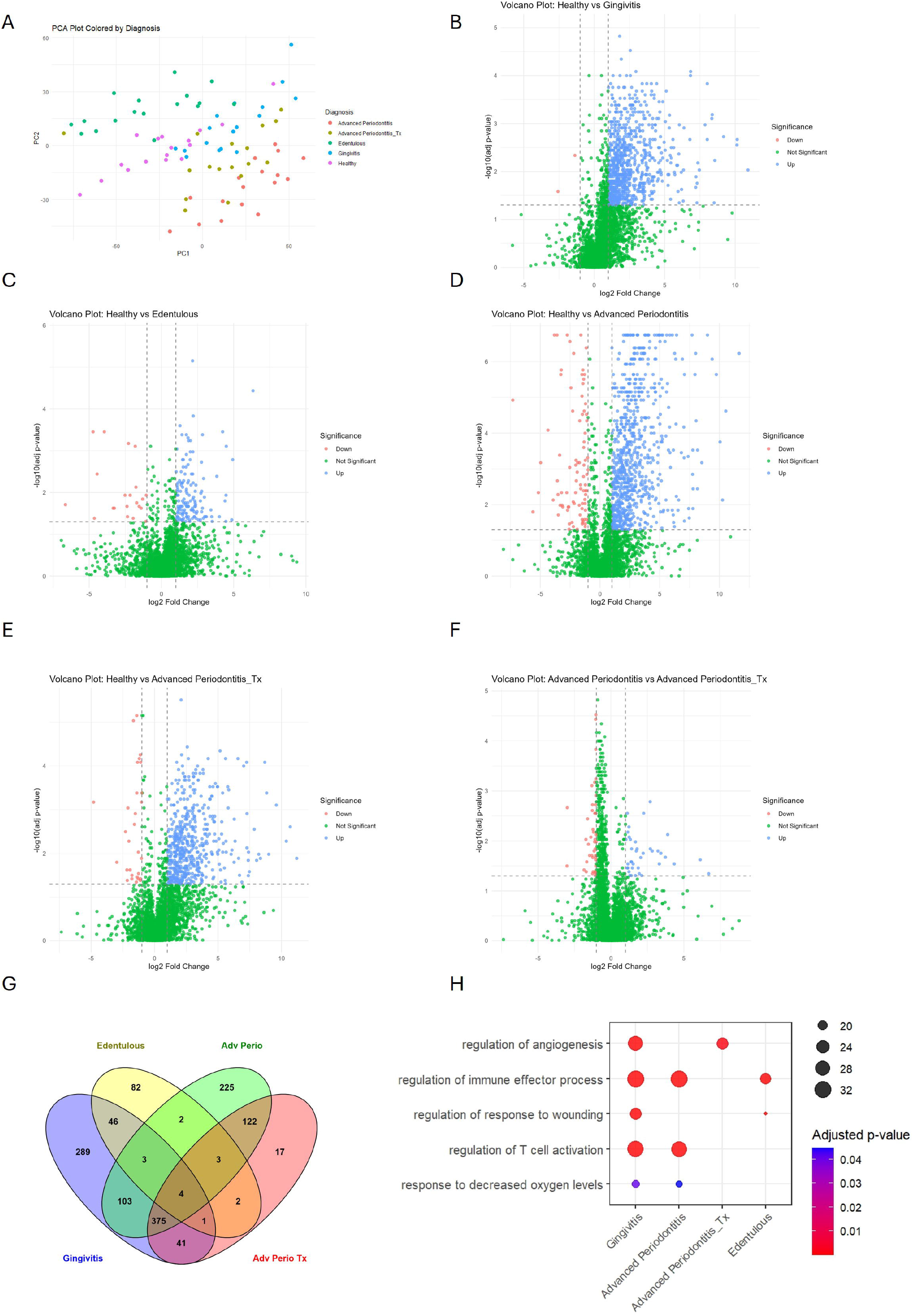
A. Principal component analysis (PCA) plot of the samples coloured by donor type. Each dot represents an individual saliva sample. B-F Volcano plots for all pairwise comparisons. Adjusted p value derived from false discovery rate adjusted p value from Mann Whitney U comparisons. Green data are not significantly (adj P value <0.05) nor greater than 2-fold changed. Red data are significantly and greater than 2-fold changed in the first group (e.g. Healthy in Healthy v Gingivitis) and Blue data significantly and greater than 2-fold changed in the second group (e.g. Gingivitis in Healthy v Gingivitis). G. Venn diagram showing comparison of proteins more abundant in the groups shown compared to health. Adv Perio = advanced Periodontitis, Tx = following periodontal treatment. H. Dot plots of gene ontology over representation for biological process for proteins increased in the volcano plots for each contrast.

**Table 3.** Numbers of proteins more (Up) or less (Down) abundant in pairwise comparisons. Increased are in group 2 eg for Healthy v Gingivitis group 2 is Gingivitis. There were greater numbers of more abundant proteins compared to less abundant proteins. The more abundant proteins were classified as to which comparison group they were significantly increased in *figure 1.

| Pairwise comparison | More abundant proteins (Up) in group 2 | Less abundant proteins (Down) in group 2 |
| --- | --- | --- |
| Healthy vs Gingivitis | 862 | 2 |
| Healthy vs Edentulous | 143 | 21 |
| Healthy vs Advanced Periodontitis | 837 | 87 |
| Healthy vs Advanced Periodontitis_Tx | 565 | 29 |
| Advanced Periodontitis vs Advanced Periodontitis_Tx | 38 | 44 |

## Discussion

The aim of this study was to evaluate for the first time differences in salivary protein overexpression between periodontal health and disease states and edentulous controls, using a high content discovery proteomics platform with high sensitivity and specificity to overcome the limitations of previously reported mass spectrometry approaches. The panel (affinity proteomics high content panel from Olink: Olink HT) analyses more than 5400 proteins and has been developed primarily for analysis of plasma or serum samples. To our knowledge it has not previously been used with saliva. The novelty and opportunity this analysis brings is to evaluate far denser data than has previously been achieved with mass spectrometry-based proteomics. Here all samples passed the QC processes and the majority of the proteins passed (only 0.3% did not meet the strict QC requirements). Forty one percent of proteins were detected above the determined LODs, similar to that reported by Rooney et al (12) for plasma proteins, though a little lower. The protein content in saliva is approximately 1 mg/ml v 70 mg/ml in plasma and whilst plasma spans 10 orders of magnitude with 99% of the proteins made up of the 12 most abundant proteins, in saliva proteins span 8 orders of magnitude and the top 20 proteins only contribute 40% of the proteome. Plasma and saliva, whilst different biofluids, share high complexity and dynamic range. The Olink HT panel has been designed to be able to cope with the high dynamic range of plasma. However, the potential to measure a large proportion Olink HT panel in saliva above the LOD offers promise for discovery science. It is also possible to use the data below the LOD for comparisons, in particular to enable the discovery of proteins that may only be present in one condition.

Further considerations include the simplicity of sample preparation and the low volumes required. In this analysis, saliva was plated and sent for analysis with no need for complex sample preparation. Equally though 50ul was required for shipping the samples to prevent sample evaporation and to allow for the automated transfer and analysis of the samples, only 2ul are required for the analysis of all 5400 proteins. This minute sample requirement means that precious samples will be available for other downstream analyses.

In this study changes to the saliva proteome were evaluated by comparing samples from healthy donors with samples from donors with periodontal diseases or donors without teeth (edentulous donors). The comparison between health and periodontal disease is one that has received a lot of interest and is well characterised, with changes in immune and inflammatory proteins acknowledged. Here we see similar patterns with changes in expected pathways (figure 1 H). This is useful as the Olink HT assay is a targeted assay and has been curated to cover the Reactome but has a heavy immune-inflammatory component. The use of samples from edentulous patients enables the view of how the teeth influence saliva composition, functioning in a similar way to a negative control. That there are many differences compared to health suggest that controlling for the number of teeth in downstream analysis could be an important factor. In this study patients needed to have at least 24 natural teeth to be included (except for the edentulous donors).

Overall, this work demonstrates that Olink HT can be used for analysis of saliva and that similar comparisons to other untargeted techniques, such as mass spectrometry-based proteomics, can be achieved.

## Data Availability

All data produced in the present study are available upon reasonable request to the authors

